# Serological Assessment of *Taenia solium* Cysticercosis Exposure in Pig Farming Population of Punjab

**DOI:** 10.1101/2025.06.11.25329407

**Authors:** Rashmi Sharma, R.S. Aulakh, Rajnish Sharma, B.B. Singh

**Affiliations:** Centre of One Health, Guru Angad Dev veterinary and Animal Sciences University, Ludhiana, 141001

## Abstract

**Background:** *Taenia solium* is a major neglected zoonotic parasite endemic to India, responsible for human cysticercosis and neurocysticercosis, particularly in pig-rearing communities. Despite increasing reports of clinical cases, limited data are available on asymptomatic exposure to *T. solium* in northern India. This study aimed to estimate the seroprevalence of *T. solium* cysticercosis and assess associated risk factors among pig-farming populations in Punjab.

**Methods:** A cross-sectional study was conducted between 2019 and 2021 in four districts of Punjab. A total of 387 individuals from pig-rearing communities were enrolled. Structured questionnaires were administered to collect demographic, occupational, and hygiene-related data. Blood samples were tested for *T. solium* IgG antibodies using a commercial ELISA kit. Fecal samples (n = 50) were also examined via sedimentation and flotation for *Taenia* eggs. Univariate and chi-square analyses were conducted to identify significant risk factors (p < 0.05).

**Results:** The overall seroprevalence of *T. solium* cysticercosis was 8.01% (31/387). The highest seropositivity was observed in Amritsar (16.9%), followed by Jalandhar (10.47%) and Ludhiana (4.58%). Pig farmers had significantly higher odds of seropositivity (11.61%) compared to non-pig farmers (4.23%) (OR = 2.97, 95% CI: 1.30 to 6.83). Seropositivity was confined to individuals with education up to 10th standard. No *T. solium* eggs were detected in any fecal sample. Significant associations were observed for location, occupation, and raw vegetable consumption. Participants who reported biannual deworming (n = 22) were all seronegative.

**Conclusions:** This study highlights ongoing exposure to *T. solium* in pig-farming communities of Punjab, with occupational and hygiene-related factors playing a critical role. Tailored interventions including regular deworming, health education, and sanitation improvements targeting high-risk groups particularly pig farmers and adults over 30 years are warranted to mitigate the transmission and burden of cysticercosis.

## Introduction

*Taenia solium* is one of the pathogenic parasites of intestinal origin. The parasite ranks first among the 24-food borne pathogen and is responsible for an economic loss of 120.7 crore in India, among which 97.8 crore in northern India [19;20;21]. Despite of that, the infection is an understated presence both in human and pig population of India. Human are the only known definitive host of parasite with neurocysticercosis being the most common infection of central nervous system of *T. solium* in human [11]. Human cysticercosis results from consumption of food and water contaminated with the eggs of *T. solium* or is observed in people who are carrier of adult parasites infections via autoinfection.

The manifestation of Cysticercosis in human is ambiguous with no particular symptom. Cases in which symptoms usually become apparent, the infection has persisted for years [5]. Numerous studies had been done in the country, but none elucidates the deciding factors for establishment of an infection with overt clinical symptoms [10].

Neurocysticercosis is the commonest cause of epilepsy worldwide [4]. The condition is the major neglected tropical zoonoses, the prevalence rate of 0.04-2.5% [16,20] have been reported in India with an unusual trait being, 95% of the cases observed in vegetarians. *Taenia solium* cysticercosis is highly prevalent in the northern region of the country, particularly in Bihar, Orissa, Uttar-Pradesh and Punjab. States of Southern India have reported low incidences of infection owing to higher literacy rates and awareness regarding the disease [8]. However, NIMHANS has reported a prevalence of 2% of neurocysticercosis in Bangalore [9].

The risk group for the disease are primarily individuals associated with pig rearing since they experience the infection first-hand through primary contact with them. In Mohanlalganj block, Lucknow, Uttar-Pradesh, a prevalence of 18.6% was stated among pig farmers with nearly 5.2% of them suffering from active epilepsy [9]. As the pig rearing industry is well established in Punjab, it calls for a need to determine the level of exposure against *T. solium* in the state. Additionally, hardly any research has been conducted in the state.

Although, cysticercosis particularly NCC is considered as a disease of low socio-economic strata, burden of inapparent infection or exposure is still unknown in larger/majority of Indian population [13]. Therefore, the following study was formulated in the pig farming community of Punjab to estimate the exposure level of *T. solium* infection in them and the predisposing risk factors.

## Methodology

### Study Area

Punjab is in northwestern region of the country and has agrarian economy with widespread livestock practices. The state spans over 50,326 square kilometers with 23 administrative districts. The state lies within the coordinates of 29.30° to 32.32° North latitude and 73.35° to 76.50° East longitude. Punjab’s rural landscape with pig farming as an emerging occupation makes it a relevant location for the present study (figure 1).

**Figure 1:**
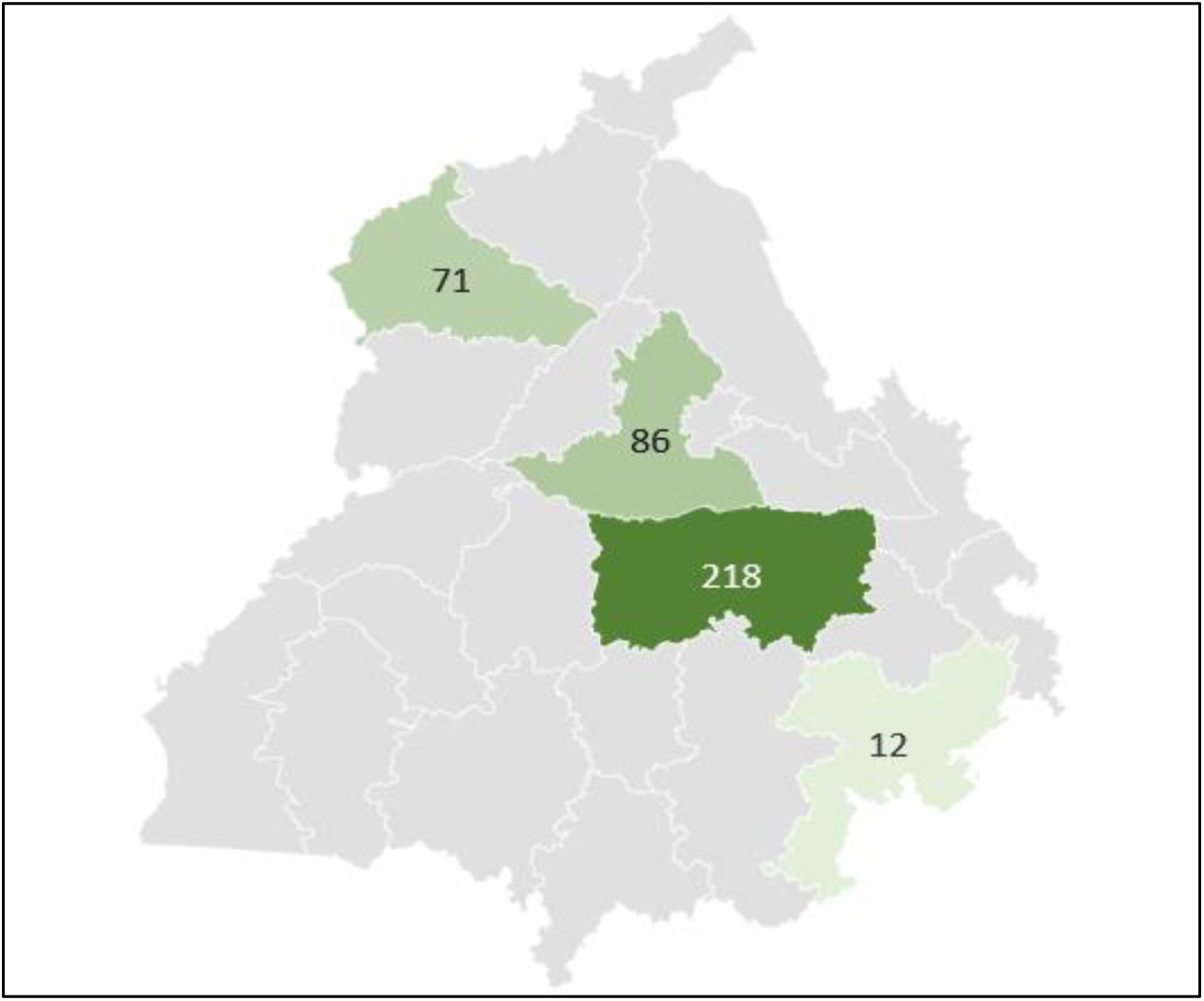
Map of Punjab from selected districts and collected sample from each district.

### Sample Size

An initial sample size of 750 was determined based on an expected response rate of 50% among the selected population for estimating the expected proportion with 5% absolute precision and 95% confidence [18]. However, due a lower-than-expected response rate, the final sample comprised of 387 participants for blood samples and 50 participants fecal samples.

### Study design

The study was conducted among various pig farming communities of selected four districts of Punjab as no one voluntarily participate from Bathinda district. The pig farming community included in the studied were - Hebbwal and Rajeev Gandhi colonies of Ludhiana, Rama mandi locality of Jalandhar, Putli Ghar area of Amritsar and a colony near bus stand in Patiala. Among the study population 51% identified pig arming as their primary occupation, while 48.8% were engaged in secondary profession like labour work and teaching. Pork and pork products were commonly accepted and consumed in the community. The study designed illustrated in figure 2.

**Figure. 2.**
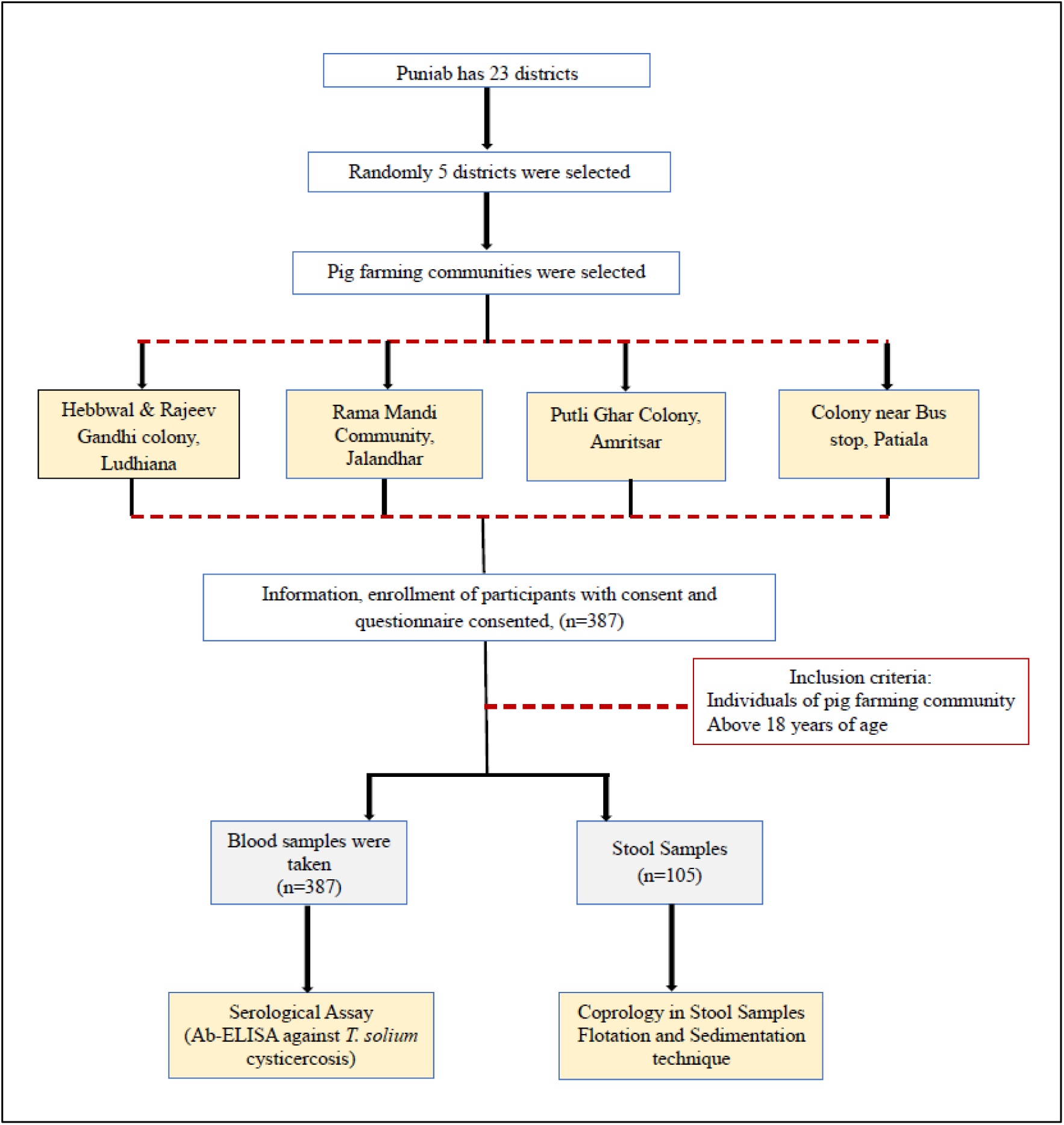
Flow Diagram of *Taenia solium* cysticercosis (serological assay) and coprology done during the study.

### Social Demographics, Behavioral and Awareness Data Collection

A structured questionnaire consisting both open and close ended questions was developed to gather relevant epidemiological data. The questionnaire focused on collecting of information related to the social demographics, pig husbandry practices and awareness of the *T. solium* cysticercosis, culinary habits and hygiene practices followed in community. Data collection was done by face-to-face interviews, during which question was administered in Hindi or English as per the preference of participant. For participants capable of reading and writing, a written version of questionnaire was provided in English and Punjabi.

### Stool Screening by Microscopy

Following the provision of written informed consent, stool samples were collected from participants in 50ml of clean and sterile containers for parasitological examination. Each container was properly labeled with the participant’s identification code to ensure traceability and confidentiality. Once collected, fresh stool samples were brought to Parasitic Zoonoses Laboratory, GADVASU. Two standard diagnostic tests employed were simple sedimentation and simple flotation technique for *Taenia* spp. ova detection.

### Blood sample collection

With written consent from participating individuals, blood sample collection was performed to carry out the serodiagnosis of *T. solium* cysticercosis. Two millitre of blood sample was collected in 5ml of venous vacutainer blood collection tube. The blood samples were centrifuges at 10000 rpm for 10 minutes to separate the serum. The extracted serum was stored in cryovials and kept at -20°C until further use.

### *T. solium* antigen detection by ELISA

Serological screening of *T. solium* cysticercosis was performed using enzyme linked immune sorbent assay (ELISA) to detect specific IgG antibodies. A commercially available ELISA kit from NovaTec was used for the purpose, following manufacturer’s instruction. The kit was pre-calibrated with standard negative and positive controls to ensure the reliability of test. In addition, known positive and negative serum samples for *Echinococcus* spp. was included in the assay to evaluate potential cross reactivity and to confirm test specificity.

The ELISA OD was read at 450nm and Ag index was calculated with the recommended cut off value: Substrate Blank: Absorbance value <0.100, negative control: Absorbance value <0.200 till Cut-off, Cut-off Control: Absorbance value -0.150-1.300 and positive Control: Absorbance value >1.300. The reported sensitivity and specificity of ELISA was 78.2%-100% and 96.67%-100% respectively. This standardized and validated approach ensured the accurate detection of seroprevalence of *T. solium* cysticercosis within the studied communities, providing critical insights into the zoonotic burden associated with pig farming practices in the region.

### Statistical analysis

Descriptive analysis of the study population was conducted. Prevalence of *Taenia solium* exposure was estimated using NADRES V_2._ A univariate analysis was conducted to determine the distribution and characteristics of Individual variable using SPSS 16 (IBM, Statistical Package for Social Sciences, version 26.0.0). The significance of various variable was assessed with p-value <0.05 as statistically significant.

### Ethical Statement

The ethical clearance to conduct the study was sort from Institutional ethics committee of Dayanand Medical College, Ludhiana, Punjab for the period of study (July 2019 till December 2021, IEC Ref No. 2020-553). All participants were informed regarding the purpose of study and voluntary written informal consent was taken before blood and stool sample collection. All procedures regarding human sample collection were in adherence to Institutes ethical conduct.

## Results

### Participant Characteristics

A total of 387 individuals from farming communities of Punjab-Amritsar (18.3%), Jalandhar (22.2%), Ludhiana (56.3%), and Patiala (3.1%) were enrolled in the study. The majority of respondents were male (74.90%) aged maximum participation was from age group upto 30 years of age (44.44%) with 18-30years. Approximately, 51.2% of the study population were actively engaged in pig farming with majority in free range (28.90%) and semi-intensive (22.20%) system of farming. Most of the participants had completed primary education upto 5^th^ standard (Table 1).

**Table 1:** Frequency and frequency percentage for *Taenia solium* infection in individuals of pig farming communities of Punjab.

| S. No. | Variable | Categories | Frequencies | Frequency Percent (%) |
| --- | --- | --- | --- | --- |
| Social Demographics Information |  |  |  |  |
| 1 | Location | Amritsar | 71 | 18.30% |
|  |  | Jalandhar | 86 | 22.20% |
|  |  | Ludhiana | 218 | 56.30% |
|  |  | Patiala | 12 | 3.10% |
| 2 | Age | 18-30 | 172 | 44.44% |
|  |  | 31-40 | 97 | 25.10% |
|  |  | 41-50 | 74 | 19.10% |
|  |  | 51-60 | 27 | 7.00% |
|  |  | >61 | 17 | 4.40% |
| 3 | Gender | Male | 290 | 74.90% |
|  |  | Female | 97 | 25.10% |
| 4 | Educational Qualification | 0-5<sup>th</sup> | 240 | 62.00% |
|  |  | 6-8<sup>th</sup> | 81 | 20.90% |
|  |  | 9-10<sup>th</sup> | 58 | 15.00% |
|  |  | 11-15 | 8 | 2.10% |
| 5 | Occupation | Pig Farmer | 198 | 51.20% |
|  |  | Not a Pig Farmer | 189 | 48.80% |
| 6 | Deworming | Yes | 22 | 94.30% |
|  |  | No | 365 | 5.70% |
| Risk factors associated |  |  |  |  |
| 7 | Have you ever heard about cysticercosis? | Yes | 15 | 3.90% |
|  |  | No | 372 | 96.10% |
| 8 | Do you the etiology of cysticercosis? | Yes | 5 | 1.30% |
|  |  | No | 382 | 98.70% |
| 9 | What leads to Infection for cysticercosis? | Know | 6 | 1.60% |
|  |  | Did not know | 381 | 98.40% |
| 10 |  | Animal | 15 | 3.90% |
|  | **Is cysticercosis disease of animal/human only or both?** | Human | 48 | 12.40% |
|  |  | No idea | 324 | 83.70% |
| 11 | **Are you aware of the symptoms encountered in cysticercosis infection?** | Yes | 19 | 4.90% |
|  |  | No | 368 | 95.10% |
| 12 | **How to prevent infection in case of cysticercosis?** | Know | 5 | 1.30% |
|  |  | Did not know | 382 | 98.70% |
| 13 | **Do you believe human are responsible for spread of this infection?** | Yes | 23 | 5.90% |
|  |  | No | 364 | 94.10% |
| 14 | **Do you think there's a need for toilet?** | Yes | 387 | 100.00% |
|  |  | No | 0.00 | 0.00% |
| 15 | **What is the source of water supply at home?** | Municipal supply | 379 | 97.90% |
|  |  | Handpump/ Others | 8 | 2.10% |
| 16 | **Are you a Vegetarian or non-vegetarian?** | Vegetarian | 121 | 31.30% |
|  |  | Non-vegetarian | 266 | 68.70% |
| 17 | **Do you consume raw vegetables?** | Yes | 334 | 86.30% |
|  |  | No | 53 | 13.70% |
| 18 | **Do you wash your vegetables before consumption?** | Yes | 387 | 100.00% |
|  |  | No | 0 | 0.00% |
| 19 | **What do you for washing hands?** | Water | 0 | 0.00% |
|  |  | Soap plus Water | 387 | 100.00% |
| 20 | **Do you wash your hands before eating?** | Yes | 387 | 100.00% |
|  |  | No | 0 | 0.00% |
| 21 | **Do you wash hands after eating?** | Yes | 387 | 100.00% |
|  |  | No | 0 | 0.00% |
| 22 |  | Yes | 387 | 100.00% |
|  | **Do you wash your hands after defecation?** | No | 0 | 0.00% |
| 23 | **What farming practice followed?** | Free range | 112 | 28.90% |
|  |  | Semi-intensive | 86 | 22.20% |
|  |  | Extensive | 8 | 2.10% |

Deworming coverage was low with only 5.70 % previously dewormed. Awareness (1.3%) regarding cysticercosis was poor with minimal recognition of symptoms (4.9%) and preventive measures (1.3%) of disease. Nearly, all participants necessitate the importance of indoor toilet’s (100%) and access to municipal water supply (97.9%), Dietary habits indicate majority of the participants are non-vegetarians (68.7%) and 86.3% of them eat properly washed raw vegetables. All participants had satisfactory hand hygiene practices with universal use of soap and water for hand washing.

### Seroprevalence of *T. solium* cysticercosis

Among 387 tested human sera samples, 31 tested positive with an overall apparent prevalence of 8.01% tested for antibodies to *T. solium* with Amritsar (16.90%) exhibiting highest true prevalence followed by Jalandhar (10.47%) and Ludhiana (4.58%; Table 2). No positive cases were reported in Patiala. A higher number of males participated in the study nearly 3 times females, with prevalence rate of 7.58% (22/290) males whereas it was 9.27% (9/97) for female. Among age groups, the 31-40year cohort had the highest seroprevalence (11.62%), followed by those aged over 60 years (12.10%). However, age was not significantly associated with IgG positivity (p = 0.550; Table 3)

**Table 2:** Apparent and true seroprevalence of *Taenia solium* in pig rearing communities in Punjab.

| Parameter | Sub-category | No. of samples collected | No. of Positive samples | Apparent Prevalence | 95% Confidence Interval (CI) | True Prevalence | Confidence interval (CI) |
| --- | --- | --- | --- | --- | --- | --- | --- |
| Location | Amritsar | 71 | 12 | 16.90% | 9.94%-27.26% | 17.87% | 10.68%-28.35% |
|  | Bathinda | 0 | 0 | 0.00% | 0.00% | 0.00% | 0.00% |
|  | Jalandhar | 86 | 9 | 10.47% | 5.17%-19.90% | 10.64% | 5.73%-18.92% |
|  | Ludhiana | 218 | 10 | 4.58% | 2.51%-8.24% | 4.03% | 2.12%-7.53% |
|  | Patiala | 12 | 0 | 0.0% | 0%-24.25% | 0.00% | 0.00% |
| Gender | Male | 290 | 22 | 7.58% | 5.07%-11.22% | 7.40% | 4.91%-11.00% |
|  | Female | 97 | 09 | 9.27% | 4.96-16.70% | 9.30% | 4.97%-16.73% |
| Age | 18-30 | 172 | 9 | 5.23% | 2.78%-9.65% | 4.75% | 2.44%-9.03% |
|  | 31-40 | 97 | 11 | 11.34% | 6.45%-19.17% | 11.62% | 0.66%-19.51% |
|  | 41-50 | 74 | 6 | 8.11% | 3.77%-16.58% | 7.99% | 6.69%-16.43% |
|  | 51-60 | 27 | 3 | 11.11% | 3.85%-28.06% | 11.36% | 3.98-28.36% |
|  | >61s | 17 | 2 | 11.77% | 3.29%-34.34% | 12.10% | 2.57%-34.46% |
| Educational Qualification | 1<sup>st</sup> -5<sup>th</sup> | 221 | 19 | 8.60% | 5.57%-13.04% | 8.54% | 5.14%-13.52% |
|  | 6<sup>th</sup> - 8<sup>th</sup> | 74 | 7 | 9.46% | 4.66%-18.26% | 9.51% | 4.11%-19.40% |
|  | 9<sup>th</sup> – 10<sup>th</sup> | 53 | 5 | 9.43% | 4.10%-20.25% | 9.48% | 3.48%-21.63% |
|  | 11<sup>th</sup>-15<sup>th</sup> | 8 | 0 | 0.00% | 0.00%-32.44% | 0.00% | 0.00%-35.53% |
| Occupation | Pig farmer | 198 | 23 | 11.61% | 7.87%-16.83% | 11.93% | 8.12%-17.18% |
|  | Not a pig farmer | 189 | 8 | 4.23% | 2.16%-8.13% | 3.63% | 1.76%-7.35% |
| Pig Farming Practices | Free-range | 112 | 13 | 11.60% | 6.91%-18.85% | 11.92% | 7.15%-19.22% |
|  | Semi-intensive | 86 | 10 | 11.62% | 6.44%-20.10% | 11.94% | 6.67%-20.47% |
|  | Intensive | 8 | 0 | 0.00% | 0.00% | 0.00% | 0.00% |

**Table 3:**
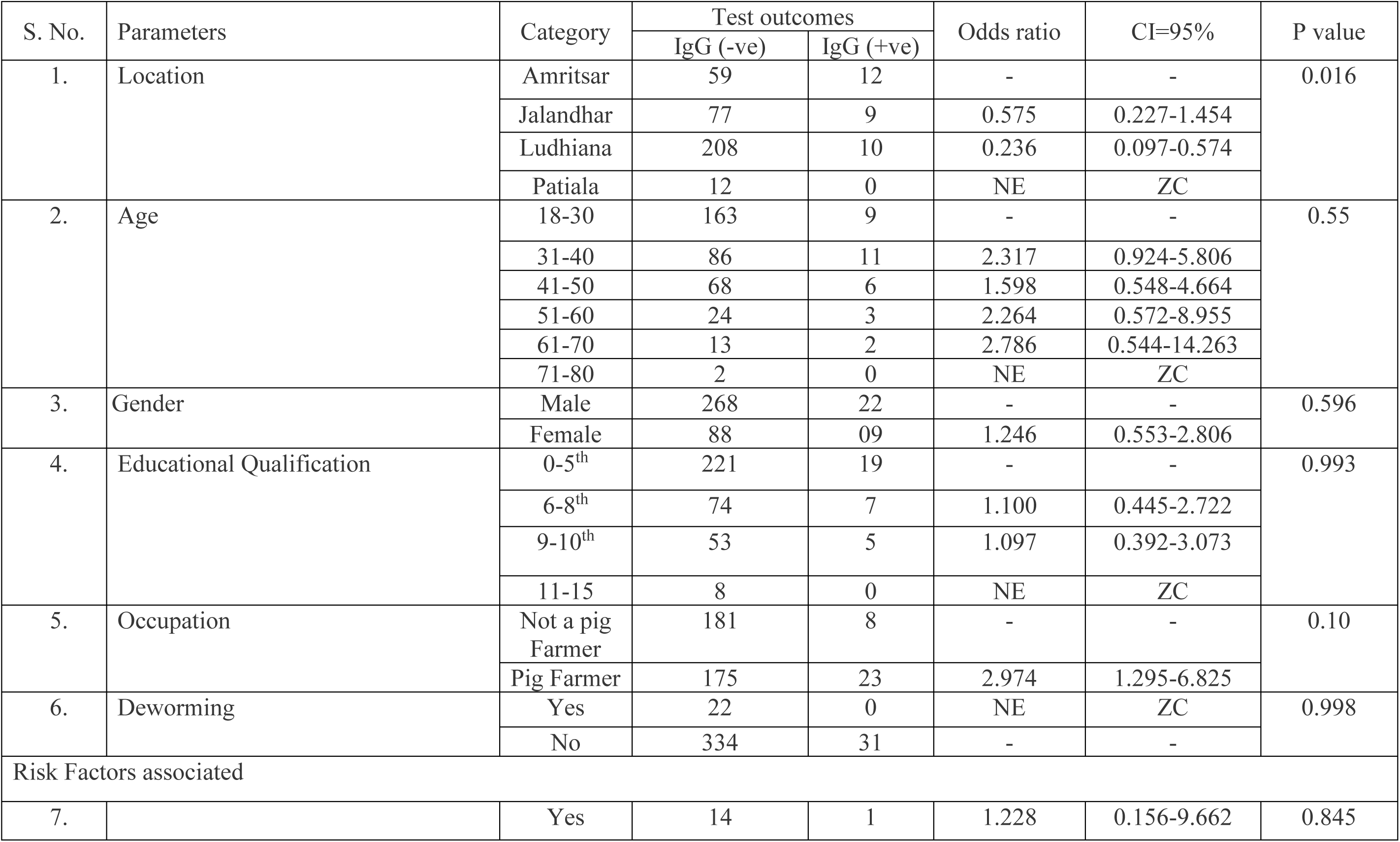
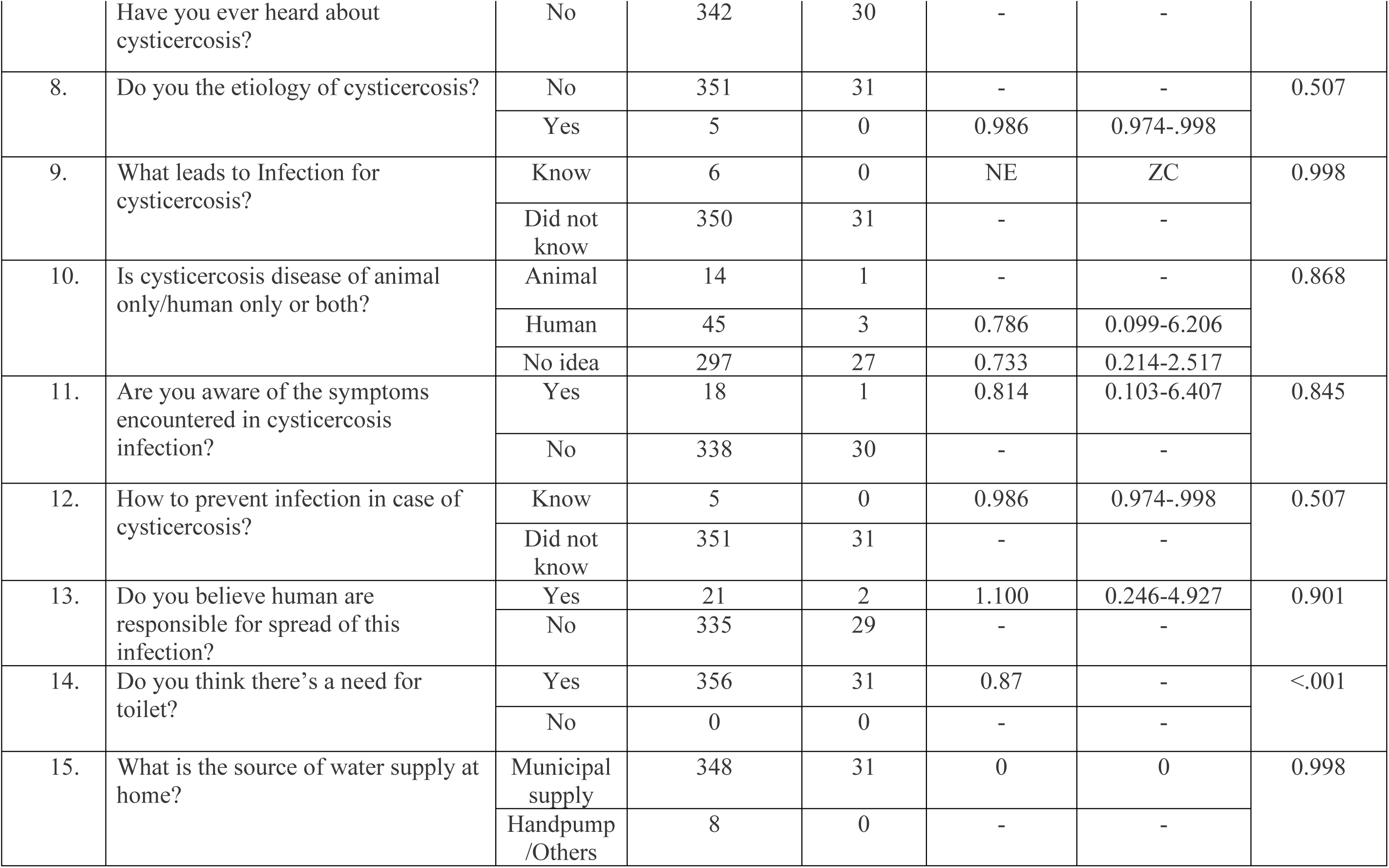
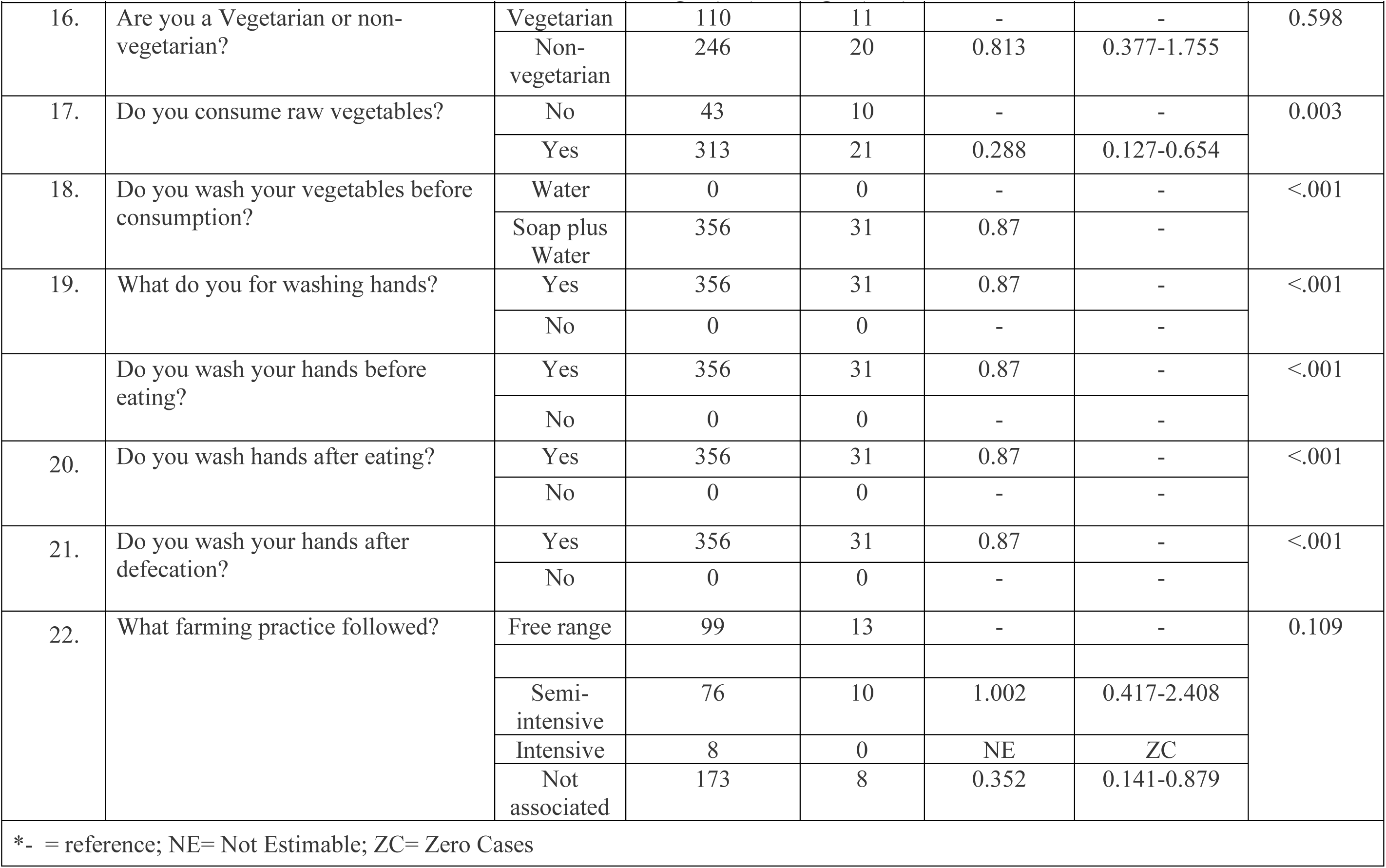
Contingency table and Chi-square test for Taenia solium infection in individuals of pig farming communities of Punjab.

Seropositivity was recorded among different education group, with highest being recorded in upto 10^th^ standard (9.43%), respectively. A substantially higher number of positive cases were recorded in participants with pig farming (11.61%; Table 2) compared to 4.23 % among not engaged in pig farming, indicating a threefold increase risk of occupational exposure. Seropositivity recorded for education group of up till 5^th^ and 10^th^ standard were 8.46% and 9.43%, respectively. Majority of seropositive cases were the individuals associated with pig farming than individuals belonging to other professions. Thus indicating, that regular deworming is an effective preventive measure in combating *T. solium* infections, however there was sample disproportion (22 vs 365) limit the strength of this association and warrants further investigation.

Coprological examination of stool samples using sedimentation and flotation methods did not reveal any *Taenia solium* eggs in any of the individuals tested, indicating no active intestinal taeniasis in the study population.

### Risk Factors Associated with Seropositivity

Geographical location (p = 0.016), occupation (p = 0.010), and raw vegetable consumption (p = 0.003) were found to have statistically significant associations with IgG seropositivity against *T. solium*. Participants from Ludhiana had significantly lower odds of infection (OR = 0.24; 95% CI: 0.10–0.57) compared to participants of Amritsar. On statistical analysis, individuals involved in pig farming were found to be three times more likely to be seropositive than non-pig farmers (OR = 2.97; 95% CI: 1.30–6.83). Interestingly, individuals who consumed raw vegetables had significantly lower odds of testing positive (OR = 0.29; 95% CI: 0.13–0.65; Table 3).

No statistically significant associations were found between *T. solium* exposure to gender, educational qualification, farming practices, and reported hygiene behaviors (including handwashing and vegetable washing practices) (p > 0.05; Table 3). Although 100% of participants reported consistent hand hygiene and vegetable washing, awareness of cysticercosis remained extremely low considering the study was done during COVID-19.

## Discussion

In the present study, we did a serological investigation in Pig farming communities of Punjab, India to estimate the *T. solium* exposure burden in the population and the risk factors associated with the disease. The overall seroprevalence was 8.01%, which is lower than that reported in several other regions of India. This could be due to low anthelminthic coverage in adult population, who are often excluded from routine mass deworming campaigns that primarily target the children. As adult are more involved in outdoor and pig rearing activities, their untreated status may contribute to sustained transmission of infection of *T. solium* in such communities [15]. Higher prevalence rates have been reported in Dehradun [32%; 8] and Vellore [19.2%; 16], which may reflect the different populations studied. Both were hospital-based studies that likely included symptomatic individuals (e.g., epilepsy patients), leading to selection bias and inflated prevalence. In contrast, this study was pig farming community-based involving asymptomatic individuals, making the results more reflective of community endemicity.

Also, the study was conducted during COVID-19, could be one of the plausible explanations for the comparatively lower prevalence observed in our study. During this period, widespread public health campaigns promoted by Indian government significantly improved hand hygiene and sanitation behavior among the general population. Given that *T. solium* transmission is fecal–oral, primarily through poor personal hygiene and contaminated food or water, these behavioral changes could have reduced incidental exposure [22; 5]. This provides a timely example of how non-targeted public health measures (e.g., handwashing promotion during a respiratory pandemic) can yield supplementary benefits in controlling enteric parasitic infections.

A significant association was observed between occupation and seropositivity. Individuals engaged in pig farming had nearly threefold higher odds of being seropositive (11.61%) compared to non-pig farmers (4.23%). This finding is justified by the closer and more frequent contact that pig farmers have with pigs and their environment, increasing the probability of exposure to *T. solium* eggs, especially in the absence of controlled sanitation and veterinary practices. Previous studies have similarly identified pig farming as an occupational risk factor for cysticercosis (14; 1].

Education level was as another influencing factor. Seropositivity was confined to individuals with education up to the 10^th^ standard, and no cases were detected in those with higher educational qualification. This supports the justification that education enhances awareness about disease transmission, hygiene, and food safety practices, thereby reducing exposure. Higher education is often linked with better socioeconomic status, access to clean water, and informed health behaviors—all protective factors against *T. solium* transmission [12; 17; 6].

Regular deworming appeared to have a protective effect; none of the individuals who underwent biannual deworming tested positive. This suggests that routine antiparasitic treatment can reduce host susceptibility or parasite burden. However, the strength of this association is limited by the small sample size in the dewormed group (n = 22), and should be interpreted cautiously. Still, the biological plausibility is strong, as deworming interrupts the lifecycle of *T. solium* and is a well-established preventive strategy [24].

Age was also a relevant factor, with individuals aged ≥31 years showing higher seropositivity (11.40%) than younger adults (6.67%). This may be explained by cumulative exposure risk over time, as older individuals typically spend more time engaged in outdoor occupational activities (including farming), increasing the likelihood of contact with contaminated sources. In contrast, younger individuals may be more protected due to school attendance, closer parental supervision, or less occupational exposure [3; 23].

The present study also observed a slightly higher seroprevalence among females (9.27%) than males (7.58%), though the difference was not statistically significant. Unlike previous studies where males were more affected [8], the reversed pattern in our findings may be contextually explained by the active role of women in household pig-rearing and food preparation in rural Punjab. These responsibilities may increase their exposure to contaminated meat or environments if proper sanitation is not followed.

Consumption of raw vegetables was significantly associated with seropositivity. This is biologically plausible, as vegetables grown in contaminated soil or irrigated with unsafe water can carry infective *T. solium* eggs. In rural settings, where open defecation or use of pig manure as fertilizer is common, the risk of foodborne cysticercosis increases. Previous studies in both children [2] and adult populations [7] have similarly reported raw vegetable consumption as a key risk factor, reinforcing the need for food safety education.

Overall, the low seroprevalence observed in this study may be justified by the combined effects of improved hygiene practices during the pandemic, shifts in dietary behavior, increased public health messaging, and possibly regional differences in environmental contamination levels. Nevertheless, the identified occupational and behavioral risk factors underscore the need for sustained One Health interventions—including pig deworming, human health education, and improved sanitation infrastructure—to reduce *T. solium* transmission in endemic rural communities.

## Conclusion

The findings of this study reaffirm that *Taenia solium* infection remains a significant concern among individuals involved in pig farming, who may serve as potential reservoirs and facilitators of transmission to household members and the broader community. The highest seroprevalence was observed in individuals aged over 30 years, suggesting that occupational exposure contributes to increased risk in this age group. Targeted public health interventions—such as awareness campaigns, educational workshops, and the distribution of informational materials focused on *T. solium* transmission, prevention, and hygiene, specifically aimed at adults engaged in pig farming, could play a pivotal role in controlling the spread of cysticercosis within endemic rural populations.

## Data Availability

All data produced in the present study are available upon reasonable request to the authors

## Acknowledgment

Rashmi Sharma (L-2018-V-21-D; 3/1/3/JRF-2018; HRD-039) gratefully acknowledges the financial and academic support provided by the Indian Council of Medical Research (ICMR), New Delhi, India.

